# Uptake of Sotrovimab for prevention of severe COVID-19 and its safety in the community in England

**DOI:** 10.1101/2022.08.17.22278893

**Authors:** Martina Patone, Andrew JHL Snelling, Holly Tibble, Carol AC Coupland, Aziz Sheikh, Julia Hippisley-Cox

## Abstract

Sotrovimab is a neutralising monoclonal antibody (nMAB), currently administrated in England to treat extremely clinically vulnerable COVID-19 patients. Trials have shown it to have mild or moderate side effects, however safety in real-world settings has not been yet evaluated. We used national databases to investigate its uptake and safety in community patients across England. We used a cohort study to describe uptake and a self-controlled case series design to evaluate the risks of 49 pre-specified suspected adverse events in the 2-28 days post-treatment. Between December 11, 2021 and May 24, 2022, there were 172,860 COVID-19 patients eligible for treatment. Of the 22,815 people who received Sotrovimab, 21,487 (94.2%) had a positive SARS-CoV-2 test and 5,999 (26.3%) were not on the eligible list. Between treated and untreated eligible individuals, the mean age (54.6, SD: 16.1 vs 54.1, SD: 18.3) and sex distribution (women: 60.9% vs 58.1%; men: 38.9% vs 41.1%) were similar. There were marked variations in uptake between ethnic groups, which was higher amongst Indian (15.0%; 95%CI 13.8, 16.3), Other Asian (13.7%; 95%CI 11.9, 15.8), White (13.4%; 95%CI 13.3, 13.6), and Bangladeshi (11.4%; 95%CI 8.8, 14.6); and lower amongst Black Caribbean individuals (6.4%; 95%CI 5.4, 7.5) and Black Africans (4.7%; 95%CI 4.1, 5.4). We found no increased risk of any of the suspected adverse events in the overall period of 2-28 days post-treatment, but an increased risk of rheumatoid arthritis (IRR 3.08, 95% CI 1.44, 6.58) and of systematic lupus erythematosus (IRR 5.15, 95% CI 1.60, 16.60) in the 2-3 days post-treatment, when we narrowed the risk period.

**Funding:** National Institute of Health Research (Grant reference 135561)

## INTRODUCTION

On December 2, 2021, the United Kingdom’s (UK) Medicines and Healthcare products Regulatory Agency (MHRA) approved the use of Sotrovimab (Xevudy) as a COVID-19 treatment for extremely clinically vulnerable patients in the community. Sotrovimab is a neutralising monoclonal antibody (nMAB) treatment [1]. nMABs are synthetic monoclonal antibodies that bind to the spike protein of SARS-CoV-2, preventing entry of the virus into the host cell and its subsequent replication. Sotrovimab is recommended for use as a treatment option in the UK for eligible, non-hospitalised adults and children (aged 12 years and above) with COVID-19 [2]. Non-hospitalised patients are eligible for treatment if: SARS-CoV-2 infection is confirmed by either reverse transcription-polymerase chain reaction (RT-PCR) testing or lateral flow test; they exhibit symptoms of COVID-19; show no signs of clinical recovery; and are members of the extremely clinically vulnerable list of patients, which have been identified through a set of clinical conditions determined by a group of experts, that are regarded as putting patients at increased risk of developing severe COVID-19 outcomes (i.e. hospitalisation or death) [3]. Sotrovimab is administered in a single intravenous infusion by a healthcare professional, to be initiated as soon as possible after diagnosis of COVID-19 and within five days of symptom onset.

Recent evidence [4, 5] has suggested that Sotrovimab significantly improves clinical outcomes in non-hospitalised patients with COVID-19 who are at high risk of severe COVID-19. A clinical trial reported that a single infusion of Sotrovimab reduced the risk of hospitalisation and death by an average of 79% in extremely clinically vulnerable adults with symptomatic COVID-19 infection [4]. In terms of safety, a small randomised controlled trial of 546 patients (n=184 assigned to Sotrovimab) found that side effects of Sotrovimab were only mild or moderate [6]. However, clinical trials involving several hundred people are likely to be underpowered and as a result, less likely to be able to detect very rare adverse events (for example, where the incidence is less than one in a thousand). Identification of any adverse effects is of considerable importance for assessing the risk-benefit balance of therapeutics and to inform clinical research and policy makers. It is also important to evaluate uptake of new therapeutic agents since there is evidence that when novel interventions are made available, they tend to exacerbate health inequalities (Inverse Equity Hypothesis) [7]. This is a crucial point given the well documented disparities in severe COVID-19 outcomes by ethnic group with higher risk of hospitalisation and mortality among some non-white groups⍰⍰ As large volumes of population data become available, it is important to evaluate the uptake and safety of Sotrovimab in real-world settings.in real-world settings.

Here, we used linked national datasets for England to report on the uptake of Sotrovimab treatment to ensure they are being used in line with guidance and there was no inequity of access or use of this therapeutic to prevent severe COVID-19 disease. Also, we investigated the safety of Sotrovimab in patients initiated in the community.

## RESULTS

### Characteristics of patients eligible for or treated with Sotrovimab

In England, between December 1, 2021 and May 24, 2022, there were 1,245,853 patients recorded as potentially eligible for Sotrovimab should they become infected with the SARS-CoV-2 virus on Pillar 2, on the national database held by NHS Digital which includes test results from the wider population (not in hospital setting). We identified 5,999 patients treated with Sotrovimab who were not on the eligible list. The characteristics for the combined 1,251,852 people who were eligible and/or treated with Sotrovimab are shown in **Table 1**. Their mean age was 60.6 years (SD: 17.5) and 54.8% (685,635) were women. Overall, 78.2% (979,195/1,251,852) were White, 2.7% (34,216/1,251,852) were Black Africans, 2.0% (24,788/1,251,852) were Indian and 1.4% (17,723/1,251,852) were Black Caribbean. 82.3% (1,029,964/1,251,852) had three or four doses of COVID-19 vaccine prior to Sotrovimab treatment or study end date, if not treated. The demographic and medical characteristics of the SARS-CoV-2 positive patients eligible for Sotrovimab treatment are shown in **Table 1**.

**Table 1.** Demographic characteristics of patients receiving Sotrovimab treatment in the community and patients who did not, amongst those eligible for treatment in the community in England between December 11, 2021 until May 24, 2022. Figures are counts (column %).

|  | Eligible for or treated with Sotrovimab | Eligible for or treated with Sotrovimab (SARS-CoV-2 positive) | Treated with Sotrovimab (SARS-CoV-2 positive) | Not treated with Sotrovimab (SARS-CoV-2 positive) |
| --- | --- | --- | --- | --- |
|  | *Counts (col %)* | *Counts (col %)* | *Counts (col %)* | *Counts (col %)* |
| **Total number of people** | 1,251,852 | 172,860 | 21,487 | 151,373 |
| **Eligible for Sotrovimab** | 1,245,853 (99.5) | 167,482 (96.9) | 16,109 (75.0) | 151,373 (100.0) |
| **SARS-CoV-2 positive** | 172,860 (13.8) | 172,860 (100.0) | 21,487 (100.0) | 151,373 (100.0) |
| **Sotrovimab treated patients** | 22,815 (1.8) | 21,487 (12.4) | 21,487 (100.0) | 0 |
| **COVID-19 symptomatic** | 66,425 (5.3) | 66,425 (38.4) | 8,089 (37.6) | 58,336 (38.5) |
| **Sex** |  |  |  |  |
| Women | 685,635 (54.8) | 100,975 (58.4) | 13,090 (60.9) | 87,885 (58.1) |
| Men | 555,369 (44.4) | 70,616 (40.9) | 8,369 (38.9) | 62,247 (41.1) |
| Not recorded | 10,848 (0.9) | 1,269 (0.7) | 28 (0.1) | 1,241 (0.8) |
| **Age** |  |  |  |  |
| Mean age (SD) | 60.6 (17.5) | 54.1 (18.1) | 54.6 (16.1) | 54.1 (18.3) |
| 12-16 years | 16,196 (1.3) | 3,743 (2.2) | 238 (1.1) | 3,505 (2.3) |
| 17-29 years | 54,585 (4.4) | 13,118 (7.6) | 1,230 (5.7) | 11,888 (7.9) |
| 30-39 years | 88,888 (7.1) | 22,240 (12.9) | 2,585 (12.0) | 19,655 (13.0) |
| 40-49 years | 134,921 (10.8) | 28,559 (16.5) | 3,887 (18.1) | 24,672 (16.3) |
| 50-59 years | 216,402 (17.3) | 34,567 (20.0) | 4,796 (22.3) | 29,771 (19.7) |
| 60-69 years | 242,880 (19.4) | 30,000 (17.4) | 4,369 (20.3) | 25,631 (16.9) |
| 70-79 years | 269,058 (21.5) | 25,193 (14.6) | 3,257 (15.2) | 21,936 (14.5) |
| 80-89 years | 136,111 (10.9) | 11,080 (6.4) | 917 (4.3) | 10,163 (6.7) |
| 90+ years | 19,323 (1.5) | 2,236 (1.3) | 97 (0.5) | 2,139 (1.4) |
| Not recorded | 73,488 (5.9) | 2,124 (1.2) | 111 (0.5) | 2,013 (1.3) |
| **Ethnicity** |  |  |  |  |
| White | 979,195 (78.2) | 137,892 (79.8) | 18,552 (86.3) | 119,340 (78.8) |
| Indian | 24,788 (2.0) | 3,184 (1.8) | 479 (2.2) | 2,705 (1.8) |
| Pakistani | 16,735 (1.3) | 1,689 (1.0) | 144 (0.7) | 1,545 (1.0) |
| Bangladeshi | 4,240 (0.3) | 473 (0.3) | 54 (0.3) | 419 (0.3) |
| Other Asian | 9,138 (0.7) | 1,201 (0.7) | 165 (0.8) | 1,036 (0.7) |
| Black Caribbean | 17,723 (1.4) | 2,064 (1.2) | 132 (0.6) | 1,932 (1.3) |
| Black African | 34,216 (2.7) | 4,191 (2.4) | 197 (0.9) | 3,994 (2.6) |
| Chinese | 2,613 (0.2) | 289 (0.2) | 43 (0.2) | 246 (0.2) |
| Other ethnic group | 28,421 (2.3) | 3,867 (2.2) | 349 (1.6) | 3,518 (2.3) |
| Ethnicity not recorded | 134,783 (10.8) | 18,010 (10.4) | 1,372 (6.4) | 16,638 (11.0) |
| **Vaccine status (prior to treatment /end of the study if not treated)** |  |  |  |  |
| No vaccine | 84,324 (6.7) | 7,743 (4.5) | 305 (1.4) | 7,438 (4.9) |
| 1 dose only | 23,295 (1.9) | 3,201 (1.9) | 206 (1.0) | 2,995 (2.0) |
| 2 doses only | 114,269 (9.1) | 17,577 (10.2) | 1,120 (5.2) | 16,457 (10.9) |
| 3 doses only | 819,307 (65.4) | 116,589 (67.4) | 14,480 (67.4) | 102,109 (67.5) |
| 4 doses only | 210,657 (16.8) | 27,750 (16.1) | 5,376 (25.0) | 22,374 (14.8) |
| **Hospital admission in the previous two years for Cardiovascular** |  |  |  |  |
| Venous thromboembolism | 17,695 (1.4) | 1,859 (1.1) | 390 (1.8) | 1,469 (1.0) |
| Immune thrombocytopenic purpura | 18,358 (1.5) | 2,003 (1.2) | 474 (2.2) | 1,529 (1.0) |
| Arterial thromboembolism | 63,382 (5.1) | 6,017 (3.5) | 882 (4.1) | 5,135 (3.4) |
| Cerebral venous thrombosis | 75 (0.0) | 15 (0.0) | 4 (0.0) | 11 (0.0) |
| Other arterial thrombosis | 2,955 (0.2) | 242 (0.1) | 41 (0.2) | 201 (0.1) |
| Myocardial infarction | 51,258 (4.1) | 4,801 (2.8) | 717 (3.3) | 4,084 (2.7) |
| Ischaemic stroke | 11,820 (0.9) | 1,197 (0.7) | 159 (0.7) | 1,038 (0.7) |
| Myocarditis | 634 (0.1) | 81 (0.0) | 18 (0.1) | 63 (0.0) |
| Pericarditis | 590 (0.0) | 83 (0.0) | 18 (0.1) | 65 (0.0) |
| Atrial fibrillation | 82,121 (6.6) | 8,006 (4.6) | 1,222 (5.7) | 6,784 (4.5) |
| Heart block | 32,678 (2.6) | 3,223 (1.9) | 483 (2.2) | 2,740 (1.8) |
| Ventricular tachycardia | 2,059 (0.2) | 223 (0.1) | 34 (0.2) | 189 (0.1) |
| Ventricular fibrillation | 482 (0.0) | 51 (0.0) | 13 (0.1) | 38 (0.0) |
| Arrhythmia | 127,781 (10.2) | 13,437 (7.8) | 2,154 (10.0) | 11,283 (7.5) |
| **Neurological** |  |  |  |  |
| Demyelinating disorders | 2,658 (0.2) | 454 (0.3) | 83 (0.4) | 371 (0.2) |
| Myasthenia | 6,825 (0.5) | 915 (0.5) | 161 (0.7) | 754 (0.5) |
| Guillain Barre syndrome | 611 (0.0) | 74 (0.0) | 14 (0.1) | 60 (0.0) |
| Encephalitis | 1,465 (0.1) | 202 (0.1) | 33 (0.2) | 169 (0.1) |
| Haemorrhagic stroke | 1,345 (0.1) | 142 (0.1) | 19 (0.1) | 123 (0.1) |
| Subarachnoid haemorrhage | 696 (0.1) | 81 (0.0) | 7 (0.0) | 74 (0.0) |
| Bell's Palsy | 2,385 (0.2) | 275 (0.2) | 42 (0.2) | 233 (0.2) |
| Multiple sclerosis | 45,371 (3.6) | 6,986 (4.0) | 1,176 (5.5) | 5,810 (3.8) |
| Optic neuritis | 1,375 (0.1) | 226 (0.1) | 47 (0.2) | 179 (0.1) |
| **Blood conditions** |  |  |  |  |
| Aplastic anaemia | 6,862 (0.5) | 726 (0.4) | 180 (0.8) | 546 (0.4) |
| Pernicious anaemia | 3,073 (0.2) | 383 (0.2) | 77 (0.4) | 306 (0.2) |
| Haemolytic anaemia | 1,816 (0.1) | 283 (0.2) | 65 (0.3) | 218 (0.1) |
| Disseminated intravascular coagulation | 82 (0.0) | 11 (0.0) | <5 | 7 (0.0) |
| ***Other conditions*** |  |  |  |  |
| Coeliac | 4,265 (0.3) | 694 (0.4) | 133 (0.6) | 561 (0.4) |
| Anaphylaxis | 822 (0.1) | 144 (0.1) | 28 (0.1) | 116 (0.1) |
| Cholangitis | 4,740 (0.4) | 620 (0.4) | 137 (0.6) | 483 (0.3) |
| Acute pancreatitis | 4,753 (0.4) | 511 (0.3) | 94 (0.4) | 417 (0.3) |
| Autoimmune hepatitis | 4,674 (0.4) | 777 (0.4) | 191 (0.9) | 586 (0.4) |
| Acute renal failure | 75,733 (6.0) | 7,286 (4.2) | 1,426 (6.6) | 5,860 (3.9) |
| Rhabdomyolysis | 1,707 (0.1) | 190 (0.1) | 25 (0.1) | 165 (0.1) |
| Acute liver failure | 5,292 (0.4) | 478 (0.3) | 96 (0.4) | 382 (0.3) |
| Jaundice | 50 (0.0) | 7 (0.0) | 1 (0.0) | 6 (0.0) |
| Angioedema | 1,064 (0.1) | 169 (0.1) | 23 (0.1) | 146 (0.1) |
| Transplant reject | 7,562 (0.6) | 1,100 (0.6) | 306 (1.4) | 794 (0.5) |
| Bullous pemphigoid | 2,657 (0.2) | 290 (0.2) | 34 (0.2) | 256 (0.2) |
| Inflame arthropathy | 33,965 (2.7) | 3,691 (2.1) | 642 (3.0) | 3,049 (2.0) |
| Rheumatoid arthritis | 42,621 (3.4) | 5,294 (3.1) | 1,191 (5.5) | 4,103 (2.7) |
| Systemic lupus erythematosus | 7,627 (0.6) | 1,356 (0.8) | 313 (1.5) | 1,043 (0.7) |
| Addison's disease | 6,789 (0.5) | 944 (0.5) | 227 (1.1) | 717 (0.5) |
| Crohn's disease | 21,584 (1.7) | 4,626 (2.7) | 783 (3.6) | 3,843 (2.5) |
| Colitis | 21,725 (1.7) | 4,357 (2.5) | 687 (3.2) | 3,670 (2.4) |
| Thyroiditis | 874 (0.1) | 165 (0.1) | 40 (0.2) | 125 (0.1) |
| Vasculitis | 15,566 (1.2) | 1,972 (1.1) | 414 (1.9) | 1,558 (1.0) |
| GI bleeding | 27,689 (2.2) | 3,014 (1.7) | 554 (2.6) | 2,460 (1.6) |

### Uptake of Sotrovimab

Of the 1,251,852 patients eligible for or treated with Sotrovimab, there were 172,860 (13.8%) patients with one or more SARS-CoV-2 positive tests between December 1, 2021 and May 24, 2022 based on Pillar 2 testing data. There were 22,815 (1.8%) patients who received treatment, and of these, 21,487 (94.1%) had a SARS-CoV-2 positive test recorded in either the Pillar 2 or the Blueteq database. Of those receiving the treatment, there were 8,089 (37.6%) patients who were recorded as having symptomatic COVID-19. The number of treated community patients per day is shown in **Figure 1**.

**Figure 1:**
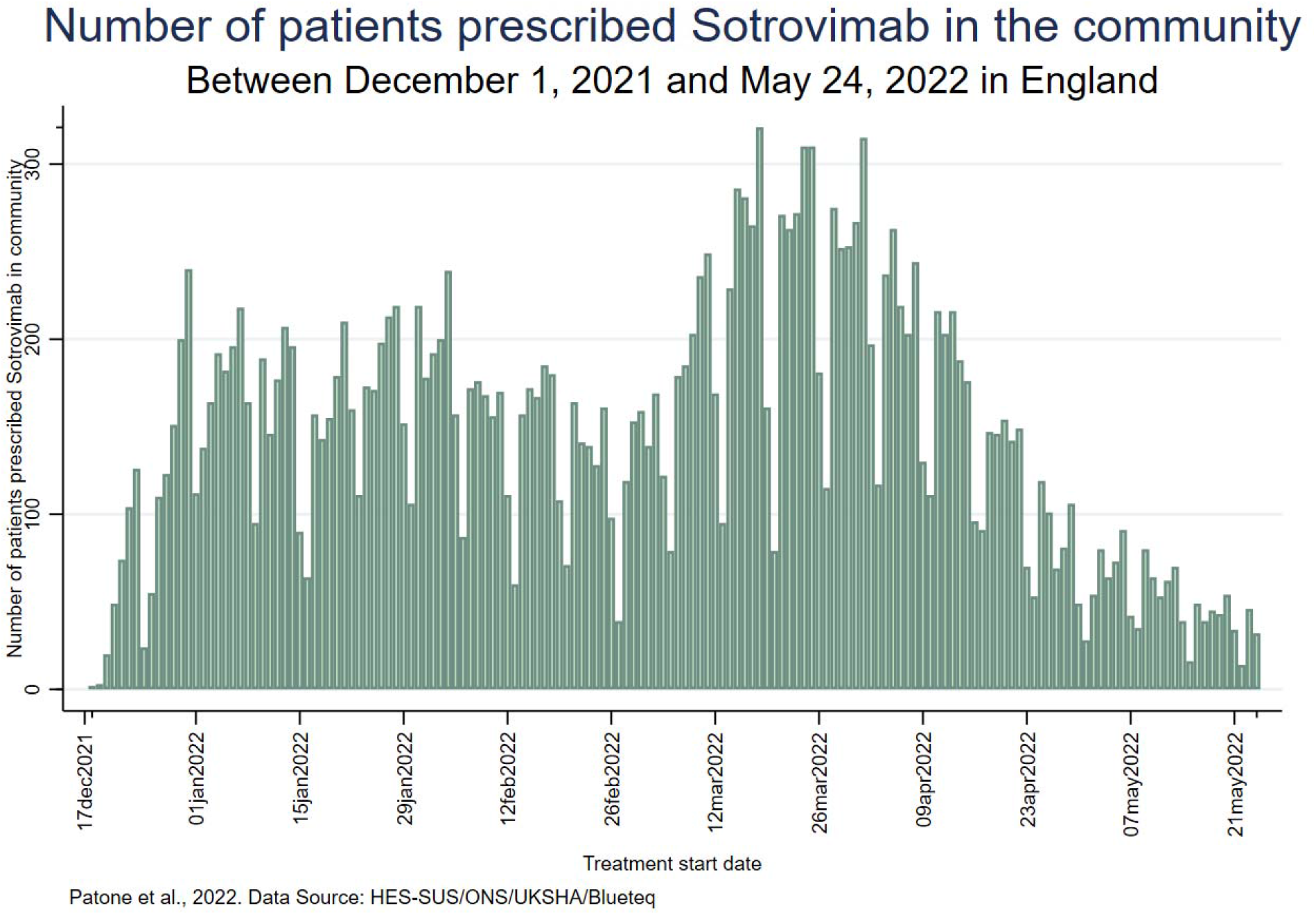
Daily number of patients prescribed Sotrovimab in the community between December 1, 2021 and May 24, 2022 in England.

We compared the demographic and medical characteristics of those treated with Sotrovimab (21,487; 12.4%) versus those who were untreated (151,373; 87.5%) within the 172,860 SARS-CoV-2 positive patients who were eligible for treatment or who had been treated. The age and sex distributions of the treated and untreated populations were similar (mean age (SD): 54.6 years (16.1) vs 54.1 years (18.3); women: 60.9% vs 58.1%).

Of those who were treated and had a SARS-CoV-2 positive test recorded, 96.3% (20,694/21,487) received the treatment in the 7 days following the test. The SARS-CoV-2 variant type had been sequenced for half (50.0%) of those who received Sotrovimab in the community. Of those, there were 6,882 (32.0%) individuals infected with the Omicron BA.1 and 3,613 (16.8%) infected with the Omicron BA.2 variant **(Supplementary Table 1)**.

Counts and proportions of SARS-CoV-2 positive patients eligible for or treated with Sotrovimab between December 11, 2021 and May 24, 2022, broken down by demographics and medical history are shown in **Table 2**. Uptake differed across ethnic groups. Higher uptake to Sotrovimab treatment was observed amongst Indian (15.0%; 95%CI 13.8, 16.3), Other Asian (13.7%; 95%CI 11.9, 15.8), White (13.4%; 95%CI 13.3, 13.6), and Bangladeshi (11.4%; 95%CI 8.8, 14.6); whereas coverage was lower amongst Black Caribbean individuals (6.4%; 95%CI 5.4, 7.5) and Black Africans (4.7%; 95%CI 4.1, 5.4).

**Table 2:** Total of SARS-CoV-2 positive community patients who received the Sotrovimab between December 11, 2021 and May 24, 2022 stratified by demographics and medical history. Figures are counts (row %).

|  | SARS-CoV-2 positive patients |  |  | SARS-CoV-2 symptomatic patients |  |  |
| --- | --- | --- | --- | --- | --- | --- |
|  | Eligible or treated with Sotrovimab | Treated with Sotrovimab | Uptake | Eligible or treated with Sotrovimab | Treated with Sotrovimab | Uptake |
|  | count | count | Row % (95%CI) | count | count | Row % (95%CI) |
| **Sex** |  |  |  |  |  |  |
| Women | 100,975 | 13,090 | 12.96 (12.76, 13.17) | 38,808 | 5,069 | 13.06 (12.73, 13.40) |
| Men | 70,616 | 8,369 | 11.85 (11.62, 12.09) | 27,149 | 3,006 | 11.07 (10.70, 11.45) |
| Not recorded | 1,269 | 28 | 2.21 (1.53, 3.18) | 468 | 14 | 2.99 (1.78, 4.99) |
| **Age** |  |  |  |  |  |  |
| 12-16 years | 3,743 | 238 | 6.36 (5.62, 7.19) | 1,476 | 94 | 6.37 (5.23, 7.73) |
| 17-29 years | 13,118 | 1,230 | 9.38 (8.89, 9.89) | 5,660 | 486 | 8.59 (7.88, 9.35) |
| 30-39 years | 22,240 | 2,585 | 11.62 (11.21, 12.05) | 9,375 | 1,075 | 11.47 (10.84, 12.13) |
| 40-49 years | 28,559 | 3,887 | 13.61 (13.22, 14.01) | 11,467 | 1,571 | 13.70 (13.08, 14.34) |
| 50-59 years | 34,567 | 4,796 | 13.87 (13.51, 14.24) | 13,812 | 1,883 | 13.63 (13.07, 14.22) |
| 60-69 years | 30,000 | 4,369 | 14.56 (14.17, 14.97) | 11,403 | 1,584 | 13.89 (13.27, 14.54) |
| 70-79 years | 25,193 | 3,257 | 12.93 (12.52, 13.35) | 8,637 | 1,073 | 12.42 (11.74, 13.14) |
| 80-89 years | 11,080 | 917 | 8.28 (7.78, 8.80) | 3,231 | 268 | 8.29 (7.39, 9.30) |
| 90+ years | 2,236 | 97 | 4.34 (3.57, 5.27) | 411 | 27 | 6.57 (4.54, 9.41) |
| Not recorded | 2,124 | 111 | 5.23 (4.36, 6.26) | 953 | 28 | 2.94 (2.04, 4.22) |
| **Ethnicity** |  |  |  |  |  |  |
| White | 137,892 | 18,552 | 13.45 (13.27, 13.64) | 52,102 | 6,920 | 13.28 (12.99, 13.58) |
| Indian | 3,184 | 479 | 15.04 (13.84, 16.33) | 1,551 | 227 | 14.64 (12.96, 16.48) |
| Pakistani | 1,689 | 144 | 8.53 (7.28, 9.96) | 933 | 77 | 8.25 (6.65, 10.20) |
| Bangladeshi | 473 | 54 | 11.42 (8.85, 14.61) | 248 | 25 | 10.08 (6.90, 14.49) |
| Other Asian | 1,201 | 165 | 13.74 (11.90, 15.80) | 531 | 68 | 12.81 (10.22, 15.93) |
| Black Caribbean | 2,064 | 132 | 6.40 (5.42, 7.54) | 855 | 36 | 4.21 (3.05, 5.78) |
| Black African | 4,191 | 197 | 4.70 (4.10, 5.38) | 1,627 | 79 | 4.86 (3.91, 6.01) |
| Chinese | 289 | 43 | 14.88 (11.22, 19.46) | 122 | 11 | 9.02 (5.06, 15.55) |
| Other ethnic group | 3,867 | 349 | 9.03 (8.16, 9.97) | 1,688 | 151 | 8.95 (7.67, 10.40) |
| Ethnicity not recorded | 18,010 | 1,372 | 7.62 (7.24, 8.01) | 6,768 | 495 | 7.31 (6.72, 7.96) |
| **Vaccine status (prior to treatment/end of the study if not treated)** |  |  |  |  |  |  |
| No vaccine | 7,743 | 305 | 3.94 (3.53, 4.40) | 3,750 | 108 | 2.88 (2.39, 3.47) |
| 1 dose | 3,201 | 206 | 6.44 (5.64, 7.34) | 1,461 | 91 | 6.23 (5.10, 7.59) |
| 2 doses | 17,577 | 1,120 | 6.37 (6.02, 6.74) | 8,227 | 502 | 6.10 (5.60, 6.64) |
| 3 doses | 116,589 | 14,480 | 12.42 (12.23, 12.61) | 43,044 | 5,728 | 13.31 (12.99, 13.63) |
| 4 doses | 27,750 | 5,376 | 19.37 (18.91, 19.84) | 9,943 | 1,660 | 16.70 (15.97, 17.44) |
| **Hospital admission**<br>**in the previous two**<br>**years for**<br>***Cardiovascular*** |  |  |  |  |  |  |
| Venous<br>thromboembolism | 1,859 | 390 | 20.98 (19.19, 22.89) | 685 | 118 | 17.23 (14.58, 20.24) |
| Immune<br>thrombocytopenic<br>purpura | 2,003 | 474 | 23.66 (21.85, 25.58) | 732 | 144 | 19.67 (16.95, 22.71) |
| Arterial<br>thromboembolism | 6,017 | 882 | 14.66 (13.79, 15.57) | 2,029 | 281 | 13.85 (12.41, 15.42) |
| Cerebral venous<br>thrombosis | 15 | <5 | 26.67 (10.38, 53.31) | 9 | <5 | 22.22 (5.60, 57.90) |
| Other arterial<br>thrombosis | 242 | 41 | 16.94 (12.72, 22.20) | 72 | 16 | 22.22 (14.08, 33.24) |
| Myocardial<br>infarction | 4,801 | 717 | 14.93 (13.95, 15.97) | 1,681 | 229 | 13.62 (12.06, 15.35) |
| Ischaemic stroke | 1,197 | 159 | 13.28 (11.48, 15.33) | 348 | 48 | 13.79 (10.55, 17.83) |
| Myocarditis | 81 | 18 | 22.22 (14.47, 32.54) | 34 | 9 | 26.47 (14.39, 43.54) |
| Pericarditis | 83 | 18 | 21.69 (14.11, 31.82) | 36 | 4 | 11.11 (4.23, 26.12) |
| Atrial fibrillation | 8,006 | 1,222 | 15.26 (14.49, 16.07) | 2,555 | 397 | 15.54 (14.18, 17.00) |
| Heart block | 3,223 | 483 | 14.99 (13.80, 16.26) | 1,061 | 158 | 14.89 (12.87, 17.16) |
| Ventricular<br>tachycardia | 223 | 34 | 15.25 (11.10, 20.58) | 96 | 14 | 14.58 (8.83, 23.13) |
| Ventricular<br>fibrillation | 51 | 13 | 25.49 (15.42, 39.11) | 23 | 6 | 26.09 (12.22, 47.23) |
| Arrhythmia | 13,437 | 2,154 | 16.03 (15.42, 16.66) | 4,584 | 734 | 16.01 (14.98, 17.10) |
| ***Neurological*** |  |  |  |  |  |  |
| Demyelinating<br>disorders | 454 | 83 | 18.28 (14.99, 22.11) | 196 | 27 | 13.78 (9.62, 19.34) |
| Myasthenia | 915 | 161 | 17.60 (15.26, 20.20) | 351 | 65 | 18.52 (14.79, 22.93) |
| Guillain Barre<br>syndrome | 74 | 14 | 18.92 (11.54, 29.45) | 27 | <5 | 14.81 (5.67, 33.46) |
| Encephalitis | 202 | 33 | 16.34 (11.85, 22.09) | 89 | 15 | 16.85 (10.42, 26.10) |
| Haemorrhagic<br>stroke | 142 | 19 | 13.38 (8.70, 20.03) | 37 | 7 | 18.92 (9.30, 34.69) |
| Subarachnoid<br>haemorrhage | 81 | 7 | 8.64 (4.18, 17.04) | 22 | <5 | 4.55 (0.64, 26.15) |
| Bell's Palsy | 275 | 42 | 15.27 (11.49, 20.02) | 120 | 26 | 21.67 (15.19, 29.92) |
| Multiple sclerosis | 6,986 | 1,176 | 16.83 (15.97, 17.73) | 2,685 | 490 | 18.25 (16.83, 19.76) |
| Optic neuritis | 226 | 47 | 20.80 (16.00, 26.58) | 95 | 11 | 11.58 (6.53, 19.71) |
| ***Blood conditions*** |  |  |  |  |  |  |
| Aplastic anaemia | 726 | 180 | 24.79 (21.79, 28.07) | 279 | 67 | 24.01 (19.36, 29.38) |
| Pernicious anaemia | 383 | 77 | 20.10 (16.39, 24.42) | 152 | 35 | 23.03 (17.02, 30.38) |
| Haemolytic anaemia | 283 | 65 | 22.97 (18.44, 28.23) | 117 | 28 | 23.93 (17.06, 32.48) |
| Disseminated<br>intravascular<br>coagulation | 11 | <5 | 36.36 (14.33, 66.12) | <5 | <5 | 66.67 (15.35, 95.66) |
| **Other conditions** |  |  |  |  |  |  |
| Coeliac | 694 | 133 | 19.16 (16.40, 22.26) | 260 | 50 | 19.23 (14.89, 24.48) |
| Anaphylaxis | 144 | 28 | 19.44 (13.78, 26.72) | 65 | 11 | 16.92 (9.63, 28.04) |
| Cholangitis | 620 | 137 | 22.10 (19.00, 25.53) | 220 | 59 | 26.82 (21.38, 33.06) |
| Acute pancreatitis | 511 | 94 | 18.40 (15.27, 21.99) | 199 | 39 | 19.60 (14.66, 25.70) |
| Autoimmune hepatitis | 777 | 191 | 24.58 (21.68, 27.73) | 311 | 90 | 28.94 (24.17, 34.23) |
| Acute renal failure | 7,286 | 1,426 | 19.57 (18.68, 20.50) | 2,383 | 467 | 19.60 (18.05, 21.24) |
| Rhabdomyolysis | 190 | 25 | 13.16 (9.05, 18.75) | 54 | 8 | 14.81 (7.59, 26.93) |
| Acute liver failure | 478 | 96 | 20.08 (16.73, 23.92) | 158 | 27 | 17.09 (11.99, 23.77) |
| Jaundice | 282 | 59 | 20.92 (16.57, 26.06) | 103 | 24 | 23.30 (16.13, 32.42) |
| Angioedema | 169 | 23 | 13.61 (9.21, 19.65) | 68 | 11 | 16.18 (9.19, 26.90) |
| Transplant reject | 1,100 | 306 | 27.82 (25.25, 30.54) | 471 | 109 | 23.14 (19.55, 27.17) |
| Bullous pemphigoid | 290 | 34 | 11.72 (8.50, 15.96) | 78 | 15 | 19.23 (11.94, 29.48) |
| Inflame arthropathy | 3,691 | 642 | 17.39 (16.20, 18.65) | 1,377 | 231 | 16.78 (14.89, 18.84) |
| Rheumatoid arthritis | 5,294 | 1,191 | 22.50 (21.39, 23.64) | 1,996 | 449 | 22.49 (20.72, 24.38) |
| Systemic lupus erythematosus | 1,356 | 313 | 23.08 (20.92, 25.40) | 595 | 126 | 21.18 (18.08, 24.64) |
| Addison's disease | 944 | 227 | 24.05 (21.43, 26.88) | 352 | 76 | 21.59 (17.60, 26.20) |
| Crohn's disease | 4,626 | 783 | 16.93 (15.87, 18.03) | 1,929 | 316 | 16.38 (14.80, 18.10) |
| Colitis | 4,357 | 687 | 15.77 (14.72, 16.88) | 1,842 | 285 | 15.47 (13.89, 17.20) |
| Thyroiditis | 165 | 40 | 24.24 (18.31, 31.36) | 72 | 18 | 25.00 (16.35, 36.24) |
| Vasculitis | 1,972 | 414 | 20.99 (19.25, 22.85) | 732 | 152 | 20.77 (17.98, 23.86) |
| GI bleeding | 3,014 | 554 | 18.38 (17.04, 19.80) | 1,101 | 196 | 17.80 (15.65, 20.17) |

Cumulative totals of SARS-CoV-2 positive eligible patients who received Sotrovimab between December 11, 2021 and May 24, 2022 stratified by demographics and medical history are shown in **Figure 2**.

**Figure 2:**
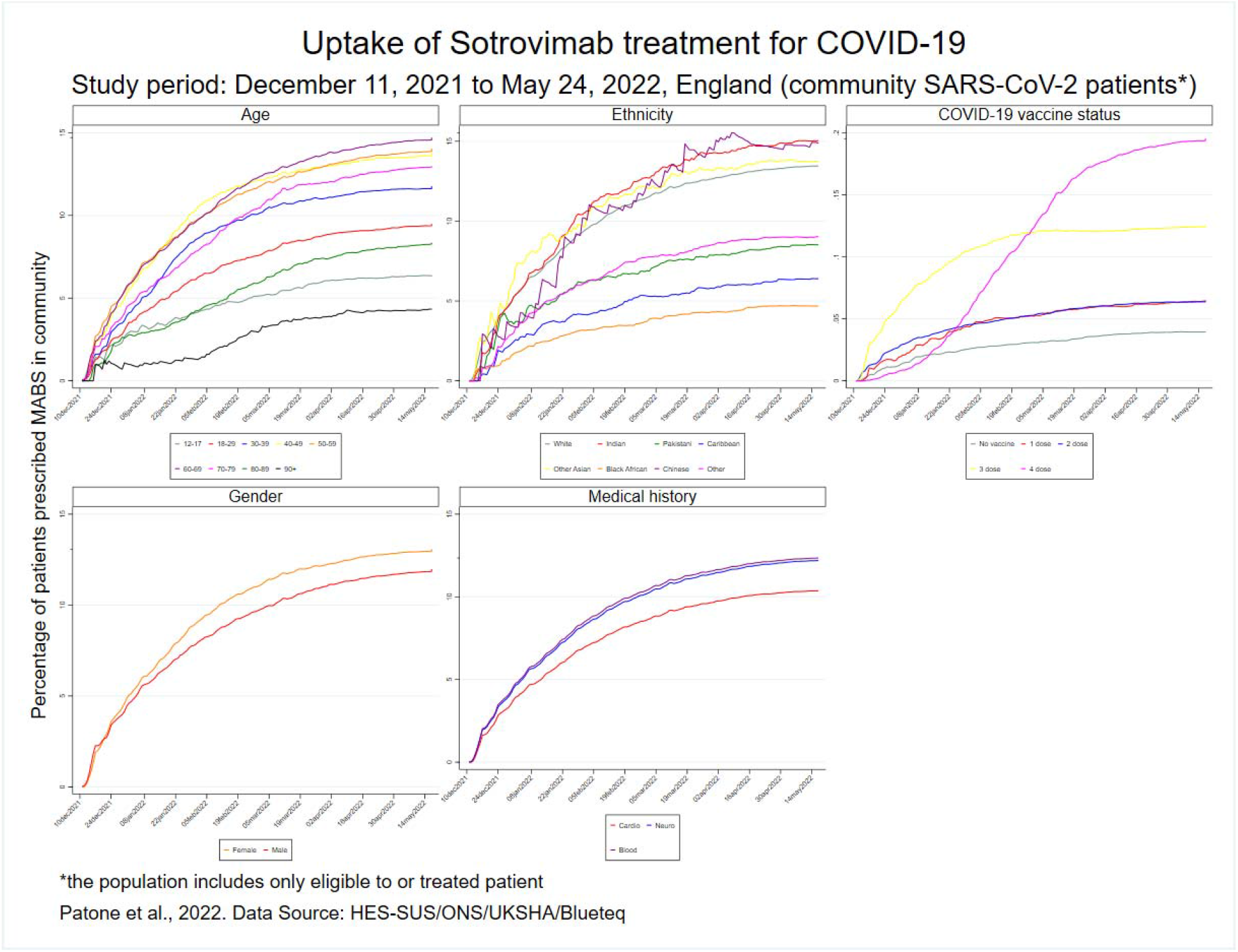
Cumulative total (%) of SARS-CoV-2 positive patients who received Sotrovimab between December 11, 2021 and May 24, 2022 stratified by demographics, vaccine status and medical history.

### Safety of Sotrovimab

Out of 49 conditions pre-specified suspected adverse events **(Supplementary Table 2)** to determine the safety of the Sotrovimab treatment, 26 conditions had at least 5 events in the 2-28 days post-treatment. Characteristics of those patients are shown in **Supplementary Table 3**.

**Table 3:** Incidence rate ratios (IRR 95% CI) for hospitalisation for individual outcomes in pre-defined risk periods immediately before and after Sotrovimab treatment, adjusted for calendar time, SARS-CoV-2 positive test and vaccine status from December 11, 2021 until May 24, 2022. The total population includes 1,251,852 patients eligible to or treated with Sotrovimab of which 22,815 were treated in the study period (cells with * are suppressed due to counts<5).

|  | **Sotrovimab** |  |  | **Sotrovimab** |  |
| --- | --- | --- | --- | --- | --- |
|  | *events* | *IRR (95% CI)* |  | *events* | *IRR (95% CI)* |
| **Venous thromboembolism** |  |  | **Autoimmune hepatitis** |  |  |
| Baseline | 3946 | 1.00 | Baseline | 575 | 1.00 |
| -28-1 days | 15 | 1.21 (0.65, 2.27) | -28-1 days | 8 | 2.35 (0.84, 6.57) |
| 0 day | <5 | n/a | 0 day | <5 | 9.75 (1.84, 51.62) |
| 1 day | <5 | 4.36 (1.53, 12.37) | 1 day | 5 | 47.76 (14.78, 154.34) |
| 2-28 days | 17 | 0.79 (0.43, 1.44) | 2-28 days | 5 | 2.04 (0.64, 6.49) |
| **Immune thrombocytopenic purpura** |  |  | **Acute renal failure** |  |  |
| Baseline | 2915 | 1.00 | Baseline | 15630 | 1.00 |
| -28-1 days | 19 | 1.18 (0.65, 2.15) | -28-1 days | 45 | 0.98 (0.69, 1.40) |
| 0 day | <5 | n/a | 0 day | <5 | 0.67 (0.17, 2.73) |
| 1 day | <5 | n/a | 1 day | 9 | 3.13 (1.58, 6.21) |
| 2-28 days | 15 | 0.81 (0.43, 1.53) | 2-28 days | 50 | 0.79 (0.56, 1.12) |
| **Arterial thromboembolism** |  |  | **Acute liver failure** |  |  |
| Baseline | 9453 | 1.00 | Baseline | 1425 | 1.00 |
| -28-1 days | 21 | 0.48 (0.27, 0.85) | -28-1 days | <5 | 1.29 (0.29, 5.62) |
| 0 day | <5 | n/a | 0 day | <5 | n/a |
| 1 day | 9 | 10.45 (4.98, 21.93) | 1 day | <5 | n/a |
| 2-28 days | 16 | 0.92 (0.50, 1.69) | 2-28 days | 6 | 1.83 (0.54, 6.18) |
| **Myocardial infarction** |  |  | **Transplant rejection** |  |  |
| Baseline | 6682 | 1.00 | Baseline | 936 | 1.00 |
| -28-1 days | 16 | 0.43 (0.22, 0.82) | -28-1 days | <5 | 0.86 (0.26, 2.82) |
| 0 day | <5 | n/a | 0 day | <5 | n/a |
| 1 day | 6 | 9.23 (3.78, 22.50) | 1 day | <5 | 5.65 (1.59, 20.10) |
| 2-28 days | 13 | 1.03 (0.53, 2.00) | 2-28 days | 5 | 0.39 (0.14, 1.10) |
| **Ischaemic stroke** |  |  | **Inflammatory arthropathy** |  |  |
| Baseline | 2975 | 1.00 | Baseline | 5112 | 1.00 |
| -28-1 days | 4 | 1.13 (0.35, 3.69) | -28-1 days | 18 | 1.17 (0.66, 2.05) |
| 0 day | <5 | n/a | 0 day | <5 | n/a |
| 1 day | <5 | n/a | 1 day | 11 | 14.42 (7.36, 28.26) |
| 2-28 days | 5 | 0.89 (0.30, 2.67) | 2-28 days | 10 | 0.57 (0.29, 1.15) |
| **Atrial fibrillation** |  |  | **Rheumatoid arthritis** |  |  |
| Baseline | 11575 | 1.00 | Baseline | 4726 | 1.00 |
| -28-1 days | 23 | 0.66 (0.41, 1.05) | -28-1 days | 37 | 1.38 (0.89, 2.13) |
| 0 day | <5 | n/a | 0 day | <5 | 1.15 (0.28, 4.78) |
| 1 day | 8 | 4.81 (2.31, 10.02) | 1 day | 35 | 26.09 (16.83, 40.45) |
| 2-28 days | 30 | 0.81 (0.53, 1.25) | 2-28 days | 23 | 0.75 (0.46, 1.24) |
| **Heart block** |  |  | **Systemic lupus erythematosus** |  |  |
| Baseline | 5883 | 1.00 | Baseline | 771 | 1.00 |
| -28-1 days | 18 | 1.01 (0.57, 1.78) | -28-1 days | 6 | 1.78 (0.61, 5.21) |
| 0 day | <5 | n/a | 0 day | <5 | n/a |
| 1 day | 5 | 6.45 (2.52, 16.51) | 1 day | 5 | 15.86 (5.50, 45.72) |
| 2-28 days | 11 | 0.60 (0.30, 1.19) | 2-28 days | 10 | 1.43 (0.61, 3.36) |
| **Arrhythmia** |  |  | **Addison's disease** |  |  |
| Baseline | 18113 | 1.00 | Baseline | 1132 | 1.00 |
| -28-1 days | 51 | 0.86 (0.62, 1.19) | -28-1 days | <5 | 0.55 (0.17, 1.79) |
| 0 day | <5 | n/a | 0 day | <5 | n/a |
| 1 day | 14 | 4.67 (2.69, 8.12) | 1 day | <5 | 20.35 (5.34, 77.55) |
| 2-28 days | 62 | 0.90 (0.66, 1.22) | 2-28 days | 5 | 1.42 (0.46, 4.33) |
| **Myasthenia** |  |  | **Crohn's disease** |  |  |
| Baseline | 766 | 1.00 | Baseline | 2087 | 1.00 |
| -28-1 days | <5 | 0.92 (0.22, 3.84) | -28-1 days | 11 | 0.92 (0.46, 1.84) |
| 0 day | <5 | n/a | 0 day | <5 | n/a |
| 1 day | <5 | 24.73 (6.43, 95.18) | 1 day | 14 | 27.07 (14.26, 51.37) |
| 2-28 days | 7 | 2.23 (0.69, 7.18) | 2-28 days | 9 | 0.71 (0.34, 1.50) |
| **Multiple sclerosis** |  |  | **Colitis** |  |  |
| Baseline | 3680 | 1.00 | Baseline | 2313 | 1.00 |
| -28-1 days | 24 | 1.63 (0.96, 2.77) | -28-1 days | 9 | 1.03 (0.47, 2.24) |
| 0 day | 5 | 3.53 (1.34, 9.30) | 0 day | <5 | n/a |
| 1 day | 37 | 42.18 (26.32, 67.60) | 1 day | <5 | 9.90 (3.41, 28.75) |
| 2-28 days | 19 | 1.01 (0.57, 1.78) | 2-28 days | 10 | 1.06 (0.50, 2.25) |
| **Aplastic anaemia** |  |  | **Vasculitis** |  |  |
| Baseline | 1193 | 1.00 | Baseline | 1678 | 1.00 |
| -28-1 days | 11 | 0.82 (0.36, 1.91) | -28-1 days | 8 | 1.05 (0.44, 2.52) |
| 0 day | <5 | n/a | 0 day | <5 | n/a |
| 1 day | <5 | 7.31 (1.62, 33.10) | 1 day | 11 | 31.71 (14.23, 70.66) |
| 2-28 days | 9 | 1.60 (0.66, 3.88) | 2-28 days | 6 | 0.71 (0.27, 1.85) |
| **Haemolytic anaemia** |  |  | **GI bleeding** |  |  |
| Baseline | 188 | 1.00 | Baseline | 5470 | 1.00 |
| -28-1 days | <5 | n/a | -28-1 days | 16 | 0.97 (0.54, 1.77) |
| 0 day | <5 | n/a | 0 day | <5 | n/a |
| 1 day | <5 | n/a | 1 day | <5 | 4.34 (1.33, 14.18) |
| 2-28 days | 5 | 1.05 (0.22, 5.06) | 2-28 days | 16 | 0.99 (0.55, 1.81) |
| **Cholangitis** |  |  |  |  |  |
| Baseline | 954 | 1.00 |  |  |  |
| -28-1 days | 5 | 0.94 (0.31, 2.85) |  |  |  |
| 0 day | <5 | n/a |  |  |  |
| 1 day | <5 | n/a |  |  |  |
| 2-28 days | 6 | 1.43 (0.49, 4.13) |  |  |  |

We found no overall association between Sotrovimab and any of the 26 suspected adverse outcomes in the 2-28 days following the treatment. Incidence rate ratios (IRRs), 95% CIs, and the number of patients with outcome events in the time period for outcomes in the overall exposure risk period of 2-28 days are shown in **Table 3** and **Figure 3**.

**Figure 3:**
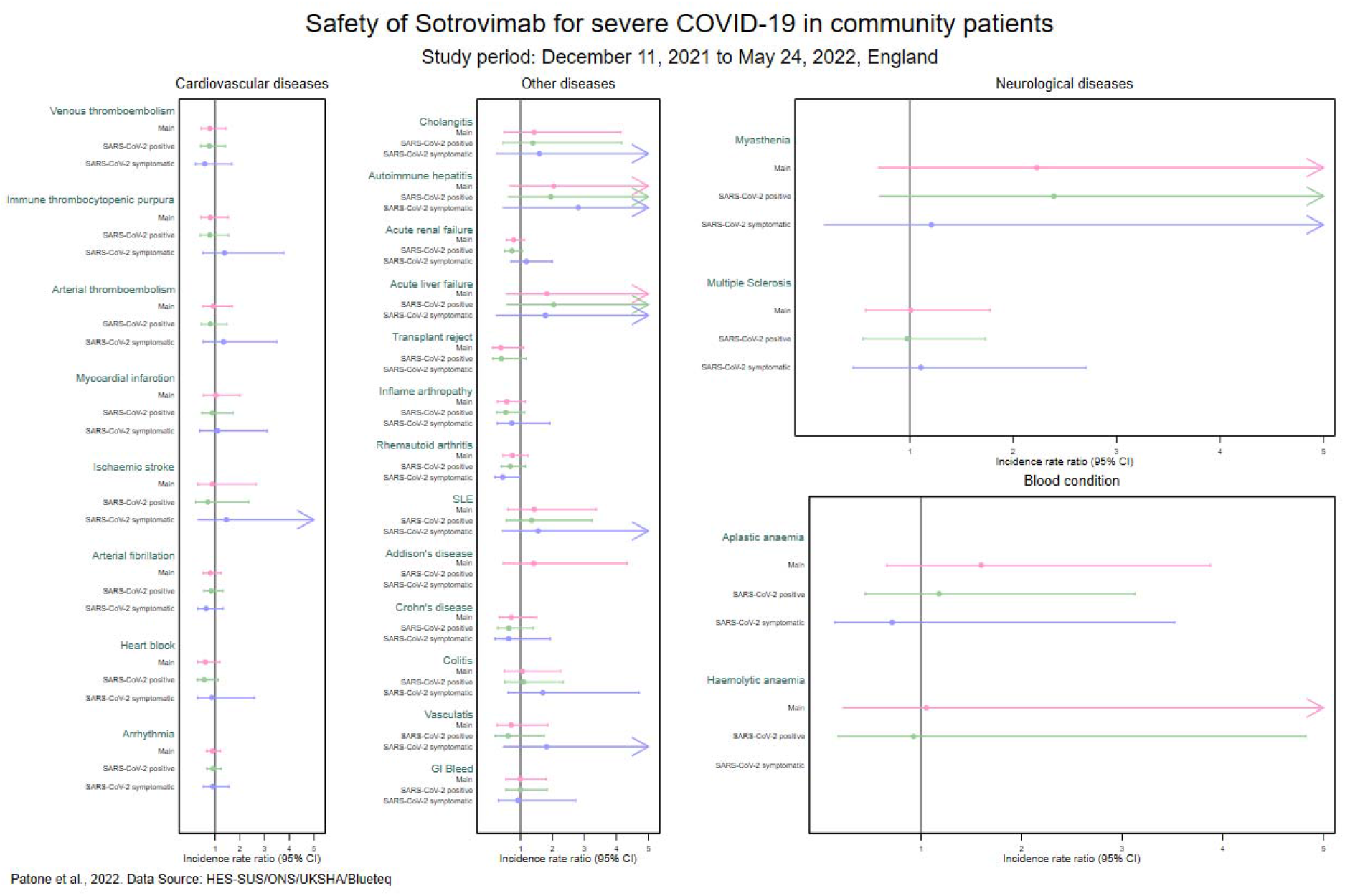
Incidence rate ratios (IRR 95% CI) for individual outcomes in the 2-28 days after Sotrovimab treatment, adjusted for calendar time, SARS-CoV-2 positive test and vaccine status, SARS-CoV-2 positive test and vaccine status from December 11, 2021 until May 24, 2022. The total population includes 1,251,852 patients eligible for or treated with Sotrovimab of which 22,815 were treated with Sotrovimab in the study period.

**Supplementary Table 4** shows the IRRs and 95% CIs and the number of patients with outcome events in the time period for outcomes in the exposure risk periods: 1-3, 4-7, 8-28 days following the treatment. Only outcomes for which we found a positive association have been reported. We found that in the 2-3 days following the treatment, there was an increased risk of admission with rheumatoid arthritis (IRR 3.08, 95% CI 1.44, 6.58) and of systematic lupus erythematosus (IRR 5.15, 95% CI 1.60, 16.60).

### Sensitivity analyses and robustness of the results

**Table 2** shows the uptake of treatments restricted only to patients recorded as having symptomatic COVID-19 disease. The difference in uptake between different ethnic groups is confirmed. In addition, uptake was lower compared to the main analysis when only COVID-19 symptomatic patients were included for the Black Caribbean (4.2%, 95% CI: 3.0, 5.8 vs 6.4%, 95% CI: 5.4, 7.5) and Chinese (9.0%, 95% CI: 5.1, 15.5 vs 14.9%, 95% CI: 11.2, 19.5) ethnic groups.).

**Supplementary Table 5 and Figure 3** compare the results of the sensitivity analyses with the main safety analysis. In both cases, when including only SARS-CoV-2 positive patients or only COVID-19 symptomatic cases, the estimates of the IRR generally agree with the main results.

## DISCUSSION

In this population-based study of more than 1 million people eligible for treatment or treated with Sotrovimab in England, we identified several key findings of policy and clinical importance.

In term of uptake, we found that about a quarter of Sotrovimab treated patients (5,999/22,815) were not on the extremely clinically vulnerable patients list and that the medical history of treated patients included co-morbidities used to identify the extremely clinically vulnerable patients, such as multiple sclerosis, acute renal failure or rheumatoid arthritis [3]. This suggests that although not every treated patient had been on the eligible list, overall, the government guidelines had been followed. Importantly, we have also found that the proportions of treated patients differed across ethnic groups, with higher uptake in White, Indian, Bangladeshi and Other Asian groups and lower in Black Caribbean and Black African groups. This suggests that some inequalities in uptake of treatment are linked to ethnicity. In terms of safety, we did not find any overall increased risk of hospitalisation for any suspected adverse events in the 2-28 days following treatment, but when splitting the 28-days risk period in narrower periods, we found an increased risk of hospital admission for rheumatoid arthritis (IRR 3.08, 95% CI 1.44, 6.58) and of systematic lupus erythematosus (IRR 5.15, 95% CI 1.60, 16.60) in the 2-3 days following the treatment.

This study had several strengths. First, we used prospectively recorded national medical data obtained from high quality, national electronic health record databases, all used for operational purposes and collected during the course of NHS clinical care giving the maximum possible power to investigate rare adverse events. The use of routinely collected electronic data means our study is not subject to recall or selection biases. Second, the breakdown of the study period in weekly blocks accounted for temporal confounding, important since the pandemic’s different waves and variants might have had different effects on health and healthcare systems. Thirdly, to assess the safety of the treatment we used the self-controlled case series (SCCS) study design, which uses a within-person comparison and hence removes potential confounding for all fixed characteristics. Also, the UK was among the first countries to roll out the COVID-19 vaccination and therapeutics treatments against COVID-19 and has some of the world’s best data for evaluating uptake and safety of new drugs [9-12]. Other strengths of our study include representativeness, data completeness and timeliness.

There were several limitations to this study. Our study evaluated clinical adverse events resulting in hospitalisation and so represents more serious effects rather than events which might occur, but not result in hospital admission. We used electronic health record data collected during the process of clinical care rather than detailed clinical assessments or questionnaire data so our focus has been on medically diagnosed conditions rather than symptoms. Although we did have data available on symptomatic SARS-CoV-2 infection, this was only recorded at a single point in time and COVID-19 symptoms may have developed over time including after the date of the test. Unfortunately, no data were available related to socio-demographics, and thus this could not be assessed as a potential confounder for associations between other factors (in particular, ethnicity) and uptake. Moreover, although we had information on all the patients treated with Sotrovimab in the community in England, out of 49 conditions studied to determine the safety of the treatment, only 26 conditions had 5 or more events in the 2-28 day period post-treatment.

The results of a descriptive cohort study of 23.4 million patients in OpenSAFELY [13] showed that between 11 December 2021 and 28 April 2022, 18,210 (18%) eligible patients with a positive SARS-CoV-2 test within the time period and registered with GP practices using the TPP database, received 1 of 5 treatments (nMABs or antivirals). Furthermore, of the 18,210 patients treated, 9,340 (51%) received treatment with Sotrovimab (9.1% of 102,170 eligible patients). Similarly, and restricting the denominator to only include eligible patients who had also received a positive SARS-CoV-2 test result or who had received treatment despite being ineligible (n=172,860), the proportion of patients who received treatment with Sotrovimab in this cohort was a comparable 12.4% (n=21,487). While the scale of the OpenSAFELY study is impressive, its treatment sample was limited to patients registered at GP practices using the TPP software (23.4 million patients). In contrast, this study utilised information for patients across the whole of England, regardless of GP practice registration. Therefore, it is likely that this study’s treatment sample is more representative. The geographic distribution of GP practices using TPP software is also problematic as it is not uniform. For example, only 17% of GP practices in London currently use TPP software. The underrepresentation of a large, ethnically heterogeneous metropolitan area such as London in the data is likely to have disproportionately impacted minority ethnic groups in OpenSAFELY’s treatment sample, as the Greater London area is the region with the largest ethnic minority population (Black and Asian) in the country, in both number (∼2.6 million) and proportion (∼ 32%) [14]. This study was not subject to this limitation.

Data on uptake of Sotrovimab in other populations internationally remains very limited. A cross-sectional analysis of nMAB (bamlanivimab; casirivimab/ imdevimab; bamlanivimab/etesevimab) uptake in the United States [15] reported that following emergency use authorisation of outpatient of nMAB treatments by the U.S. Food and Drug Administration (FDA) on November 9, 2020 and up to April 11, 2021, 69,377 patients received an infusion, nationwide. The study in question did not determine the size of the population, which may have been eligible for treatment. However, the demographic characteristics of the treated population in the US were similar to those reported in this study. The treated population were more likely to be under 65 years old (57.5%), female (53.8%), and White (54.8%). The reasons for low uptake of nMAB treatments in our study and the US, and elsewhere may be due to factors such as stockpiling, limited and/or unequal distribution of nMAB treatments throughout healthcare systems, barriers to patient uptake of treatments which require intravenous infusions, or hesitancy to adopt treatments on the part of patients or providers [15].

The inequality of uptake of novel COVID-19 treatment across ethnic groups has major implications for healthcare policy. Earliest studies [8] have showed marked disparities by ethnic group in mortality for non-white ethnic groups compared with white, and ethic differences in treatment uptake could exacerbate existing health inequalities. This information can provide evidence to enable policy makers to identify strategies to improve and strengthen the use of Sotrovimab by supporting the construction and use of targeted interventions to address these inequalities. Moreover, we found that Sotrovimab treatment is overall safe in terms of the outcomes studied, which can help to inform clinical decision making. However, these findings would benefit from corroboration from other countries using similarly robust analytical approaches and large datasets.

In summary, in this first study of safety of Sotrovimab, we have found no safety signals of concern with no increased risk of possible adverse events in the overall period of 2-28 days post treatment, but we did find an increased risk of rheumatoid arthritis and of systematic lupus erythematosus in the 2-3 days post-treatment when we narrowed the risk periods. We have, however, found evidence for inequalities in the uptake of the treatment across ethnic groups, with uptake of treatment being higher in White, Indian, Bangladeshi and Other Asian patients and lower in Black Caribbean and Black African patients. This is particularly important given existing evidence of ethnic disparities in severe COVID-19 outcomes, indicating that those who may have the greatest clinical need are not receiving treatments from which they could benefit, thereby potentially exacerbating health inequalities [7].

## Data Availability

The data that support the findings of this study Blueteq, National Immunisation (NIMS) Database of COVID-19, mortality (ONS), hospital admissions (HES) and SARS-CoV-2 infection data (PHE), are not publicly available because they are based on de-identified national clinical records. Due to national and organisational data privacy regulations, individual-level data such as those used for this study cannot be shared openly.

## ACKNOWLEDGEMENTS

This project involves data derived from patient-level information collected by the NHS, as part of the care and support of cancer patients. The SARS-Cov-2 test data are collated, maintained and quality assured by Public Health England (PHE). Access to the data was facilitated by the PHE Office for Data Release. The Hospital Episode Statistics, Secondary Users Service (SUS-PLUS) datasets and civil registration data are used by permission from NHS Digital who retain the copyright in that data. We acknowledge the contribution of EMIS practices who contribute to QResearch^®^ and EMIS Health and the Universities of Nottingham and Oxford for expertise in establishing, developing or supporting the QResearch database. NHS Digital and Public Health England bears no responsibility for the analysis or interpretation of the data. The investigators acknowledge the philanthropic support of the donors to the University of Oxford’s COVID-19 Research Response Fund

## AUTHOR CONTRIBUTIONS

MP, JHC, CC led the study conceptualisation, development of the research question. JHC obtained funding, designed the analysis, obtained data approvals, contributed to interpretation of the analysis. MP undertook the data specification, curation, analysis and wrote the first draft of the paper. HT, AS, AS contributed to the discussion on protocol development and provided critical feedback on drafts of the manuscript. All authors approved the protocol, contributed to the critical revision of the manuscript and approved the final version of the manuscript.

## METHODS

### Ethical approval

National Health Service Research Ethics Committee (NHS REC) approval was obtained from East Midlands-Derby Research Ethics Committee [reference 04/03/2021].

### Data

We used the national specialised commissioning database for England (also known as Blueteq), which includes information submitted by NHS hospitals and COVID Medicine Delivery Units (CMDUs) treating patients when an individual is determined to be clinically eligible for, and agrees to, treatment to prevent severe COVID-19. This includes the type, e.g., antivirals or nMAB including Sotrovimab, date of treatment, date of SARS-CoV-2 positive test and whether the treatment had been administrated to a community or hospital-based patient.

We linked the Blueteq database at individual level to the list of extremely clinically vulnerable patients eligible for COVID-19 treatments [3] (as well as the SARS-CoV-2 infection data (Pillar2 data), hospital admissions (Hospital Episode Statistics - Secondary Uses Service), national data for mortality (office of national statistics) and the National Immunisation (NIMS) database of COVID-19 vaccination.

### Study design

A descriptive cohort study was used to investigate uptake of Sotrovimab treatment.

The SCCS design was used to study safety of Sotrovimab. This design was originally developed to examine vaccine safety [16, 17] and has been frequently used for pharmacological vigilance [18]. The analyses are conditional on each case (person with an outcome of interest), so any fixed characteristics during the study period, such as sex, ethnicity or chronic conditions, are inherently controlled for. Age was considered as a fixed variable because the study period was short. Any time-varying factors, like seasonal variation, still however need to be adjusted for in the analyses.

### Study period and population

The study period was between December 11, 2021 to May 24, 2022 (last data update).

The study population for the cohort study included all patients recorded in the extremely clinically vulnerable patients eligible for COVID-19 treatments cohort or treated with Sotrovimab in the community in England in the study period. Patients were excluded if they had missing NHS number, were treated with antivirals, their treatment was not approved by the commissioning team or completed, tested positive in the hospital setting or were aged 12 or less. A data flow diagram is included in **Figure 1**.

We identified SARS-CoV-2 positive patients in the community in this period using Pillar 2 testing data, which includes test results in the wider population (not in hospital setting), through commercial partnerships, either processed in a lab or more rapidly via antigen tests [19].

For the safety analyses, as the SCCS method requires just the use of cases in the model, i.e. patients who died or were admitted to hospital for the specific outcome. We developed for each possible adverse event (listed in **Supplementary table 1**) a cohort consisting of patients in the study population that were admitted to hospital or died from each outcome of interest during the study period. Patients were followed up from the start (December 11, 2021) to the end of the study period or when they died. Patients who were admitted to hospital for the outcome in the two years prior study start were excluded.

### Exposures

For the safety analysis, the exposure variable was Sotrovimab treatment in a community setting, recorded in the Blueteq dataset during the study period in England.

We defined the exposure risk intervals as the 2-28 days after the Sotrovimab treatment. As a secondary aim, we also looked at the following pre-specified time-periods: 2-3, 4-7 and 8-28 days after treatment.

Since there might be a recording delay of 1 day between Blueteq and SUS-HES data (i.e. between the day of hospital admission and the day of the Sotrovimab treatment administration), results for the day following a Sotrovimab exposure could be misleading so day 0 (treatment day) and day 1 (the day after the treatment) were kept as a separate risk interval.

A pre-risk interval of 1-28 days before the Sotrovimab exposure date was included to account for potential bias that might arise if the occurrence of the outcome temporarily influenced the likelihood of exposure. The baseline period for each exposure comprised the remaining time from December 11, 2021 until 29 days before the exposure and from 29 days after the exposure until May 24, 2022 or the censored date, if earlier

### Outcomes

The outcomes of the safety analysis were selected complications, including those used in drug safety studies by U.S. Food and Drug Administration (FDA), European Medicines Agency (EMA) or MHRA and those with prior indications of association with SARS-CoV-2 infection (**see Supplementary table 1**). These outcomes were identified using all the diagnostic (International Classification of Diseases, ICD-10) codes recorded at hospital admission (13 in the SUS database and 21 in the HES database). ICD-10 codes used to identify the outcomes of interest are available at https://www.qresearch.org/data/qcode-group-library/. The outcomes were identified as the first hospital admission due to the event of interest or death recorded on the death certificate with the ICD-10 code related to the outcome of interest within the study period. Patients with community prescribed Sotrovimab may have these administered in a hospital setting. In this case, some outcomes of interest might have been recorded in the admission field if existing condition of the patient and this would make it difficult to determine the sequence of exposure and outcome. For this reason, outcomes recorded on the same day of the Sotrovimab treatment were excluded. Therefore, we defined as an outcome the earliest hospital admission in the study period which does not include the Sotrovimab treatment recorded in the same admission in the Blueteq database.

Similarly, we identified hospital admissions for each outcome in the two years prior study start (December 11, 2021) and excluded them.

### Confounders

We included in each model a first, second, third or fourth dose of the three main COVID-19 vaccines in use in the UK (I.e., ChAdOx1, BNT162b2 or mRNA-1273) in the study period. Due to the small number of events, we have not distinguished the vaccine type. We also included a SARS-CoV-2 positive test recorded in the Pillar 2 data or the Blueteq database.

Hospital admissions were likely influenced by the pressure on the health systems due to COVID-19, which was not uniform during the pandemic period. To allow for these underlying seasonal effects, we split the study observation period into weeks and adjusted for week as a factor variable in the statistical models.

### Statistical analysis

We described characteristics of the cohort eligible for or treated with Sotrovimab overall and restricted to only those positive to SARS-CoV-2 in terms of age, sex, ethnicity, vaccine status and prior medical history, and compared these in those with and without Sotrovimab treatment.

For the safety analyses we described characteristics of each set of cases (patients with the outcomes of interest) in terms of age, sex, prior medical history, by treatment. The SCCS models were fitted using a conditional Poisson regression model with an offset for the length of the exposure risk period. Incidence rate ratios (IRR), the relative rate of hospital admissions or deaths due to each outcome of interest in exposure risk periods relative to baseline periods, and their 95% CI were estimated by the SCCS model adjusted for week, positive SARS-CoV-2 test and COVID-19 vaccine dose as time-varying covariates. Sotrovimab treatment, a SARS-CoV-2 positive test, vaccine dose and calendar week were included in the same model. Separate analyses were carried out in cases with each outcome of interest. Our sample size calculations showed that data on at least 73 events were needed in people treated with Sotrovimab in the community (incl. those occurring at baseline period) to detect an IRR of 2 or more, with a significance level of 0.05 and a power of 0.80. Out of 49 conditions studied (**Supplementary Table 2**), 26 had 5 or more number of events in the 2-28 days post treatment. We have conducted analysis only for these 26 outcomes. The cut-off of 5 events has been determined in line with disclosure control policies designed to protect patient confidentiality.

We conducted sensitivity analyses restricted to only SARS-CoV-2 positive patients and COVID-19 symptomatic patients.

Stata version 17 was used for these analyses.

### Data availability

The data that support the findings of this study – Blueteq, National Immunisation (NIMS) Database of COVID-19, mortality (ONS), hospital admissions (HES) and SARS-CoV-2 infection data (PHE) - are not publicly available because they are based on de-identified national clinical records. Due to national and organisational data privacy regulations, individual-level data such as those used for this study cannot be shared openly.

### Code availability

A template code used for this study is available at https://github.com/qresearchcode/COVID-19-Treatment

## References

1. GOV.UK, MHRA approves Xevudy (sotrovimab), a COVID-19 treatment found to cut hospitalisation and death by 79%. 2022.

2. England, N., Interim clinical commissioning policy: neutralising monoclonal antibodies or antivirals for non-hospitalised patients with COVID-19. 2022.

3. GOV.UK, Highest-risk patients eligible for COVID-19 treatments: guide for patients. 2022.

4. Gupta, A., et al., Early Treatment for Covid-19 with SARS-CoV-2 Neutralizing Antibody Sotrovimab. N Engl J Med, 2021. 385(21): p. 1941–1950.

5. Gupta, A., et al., Effect of Sotrovimab on Hospitalization or Death Among High-risk Patients With Mild to Moderate COVID-19: A Randomized Clinical Trial. JAMA, 2022. 327(13): p. 1236–1246.

6. Group, A.-T.f.I.w.C.-T.S., Efficacy and safety of two neutralising monoclonal antibody therapies, sotrovimab and BRII-196 plus BRII-198, for adults hospitalised with COVID-19 (TICO): a randomised controlled trial. Lancet Infect Dis, 2022. 22(5): p. 622–635.

7. Victora, C.G., et al., The Inverse Equity Hypothesis: Analyses of Institutional Deliveries in 286 National Surveys. Am J Public Health, 2018. 108(4): p. 464–471.

8. Clift, A.K., et al., Living risk prediction algorithm (QCOVID) for risk of hospital admission and mortality from coronavirus 19 in adults: national derivation and validation cohort study. BMJ, 2020. 371: p. m3731.

9. Hippisley-Cox, J., et al., Risk of thrombocytopenia and thromboembolism after covid-19 vaccination and SARS-CoV-2 positive testing: self-controlled case series study. BMJ, 2021. 374: p. n1931.

10. Patone, M., et al., Neurological complications after first dose of COVID-19 vaccines and SARS-CoV-2 infection. Nat Med, 2021.

11. Patone, M., et al., Risk of myocarditis following sequential COVID-19 vaccinations by age and sex. medRxiv, 2021: p. 2021.12.23.21268276.

12. Patone, M., et al., Risks of myocarditis, pericarditis, and cardiac arrhythmias associated with COVID-19 vaccination or SARS-CoV-2 infection. Nat Med, 2021.

13. Green, A., et al., Trends, variation and clinical characteristics of recipients of antivirals and neutralising monoclonal antibodies for non-hospitalised COVID-19: a descriptive cohort study of 23.4 million people in OpenSAFELY. medRxiv, 2022: p. 2022.03.07.22272026.

14. GOV.UK, Regional ethnic diversity. 2020.

15. Anderson, T.S., et al., Uptake of Outpatient Monoclonal Antibody Treatments for COVID-19 in the United States: a Cross-Sectional Analysis. J Gen Intern Med, 2021. 36(12): p. 3922–3924.

16. Farrington, C.P., et al., Self-Controlled Case Series Analysis With Event-Dependent Observation Periods. Journal of the American Statistical Association, 2011. 106(494): p. 417–426.

17. Petersen, I., I. Douglas, and H. Whitaker, Self controlled case series methods: an alternative to standard epidemiological study designs. BMJ, 2016. 354: p. i4515.

18. Grosso, A., et al., Use of the self-controlled case series method in drug safety assessment. Expert Opin Drug Saf, 2011. 10(3): p. 337–40.

19. GOV.UK, NHS Test and Trace statistics (England): methodology.

