## Supplementary Tables 1-5 and supplementary Figure 1 for "Uptake of Sotrovimab for prevention of severe COVID-19 and its safety in the community in England"

**Supplementary Table 1. Additional characteristics of patients receiving the Sotrovimab treatments in the community in England between December 11, 2021 until May 24, 2022. Figures are column % (counts).**

|  | **Sotrovimab**  **(SARS-CoV-2 positive)** | **Sotrovimab**  **(SARS-CoV-2 symptomatic)** |
| --- | --- | --- |
|  | *Counts (col %)* | *Counts (col %)* |
| **Lag between positive test and treatment** |  |  |
| 0 day | 811 (3.8) | 156 (1.9) |
| 1 day | 4079 (19.0) | 1363 (16.9) |
| 2-3 days | 6887 (32.1) | 2847 (35.2) |
| 3-4 days | 5101 (23.7) | 2126 (26.3) |
| 5-7 days | 3816 (17.8) | 1345 (16.6) |
| 8-28 days | 98 (0.5) | 27 (0.3) |
| 29+ days | 321 (1.5) | 123 (1.5) |
| Treatment before positive test | 374 (1.7) | 102 (1.3) |
| **SARS-CoV-2 variant** |  |  |
| Delta | 183 (0.9) | 74 (0.9) |
| Delta plus | 17 (0.1) | 9 (0.1) |
| Omicron BA.1 | 6882 (32.0) | 3543 (43.8) |
| Omicron BA.2 | 3613 (16.8) | 1606 (19.9) |
| Unknown | 10748 (50.0) | 2835 (35.0) |

**Supplementary Table 2: List of conditions used for the safety analysis. ICD-10 codes used to identify the outcomes of interest are available at** [**https://www.qresearch.org/data/qcode-group-library/**](https://www.qresearch.org/data/qcode-group-library/)

| ***Cardiovascular*** | ***Blood conditions*** |
| --- | --- |
| Venous thromboembolism | Aplastic anaemia |
| Immune thrombocytopenic purpura | Pernicious anaemia |
| Arterial thromboembolism | Haemolytic anaemia |
| Cerebral venous thrombosis | Disseminated intravascular coagulation |
| Other arterial thrombosis |  |
| Myocardial infarction | ***Other conditions*** |
| Ischaemic stroke | Coeliac |
| Myocarditis | Anaphylaxis |
| Pericarditis | Cholangitis |
| Atrial fibrillation | Acute pancreatitis |
| Heart block | Autoimmune hepatitis |
| Ventricular tachycardia | Acute renal failure |
| Ventricular fibrillation | Rhabdomyolysis |
| Arrhythmia | Acute liver failure |
|  | Jaundice |
| ***Neurological*** | Angioedema |
| Demyelinating disorders | Transplant reject |
| Myasthenia | Bullous pemphigoid |
| Guillain Barre syndrome | Inflame arthropathy |
| Encephalitis | Rheumatoid arthritis |
| Haemorrhagic stroke | Systemic lupus erythematosus |
| Subarachnoid haemorrhage | Addison's disease |
| Bell’s Palsy | Crohn's disease |
| Multiple sclerosis | Colitis |
| Optic neuritis | Thyroiditis |
|  | Vasculitis |
|  | GI bleeding |

**Supplementary Table 3: Demographic characteristics of patients who experienced the individual outcomes in the baseline and in the 2-28 days following the Sotrovimab treatment. The total population includes 1,251,852 patients eligible to or treated with Sotrovimab of which 22,815 were treated in the study period. Out of 49 outcomes screened for safety signal, there were 26 outcomes with at least 5 events in the 2-28 days following nMAB (cells with less than 5 events are suppressed).**

|  | **Total** | | **Women** | | **Men** | | **Age** | | **Deaths** | |
| --- | --- | --- | --- | --- | --- | --- | --- | --- | --- | --- |
|  | **Baseline** | **2-28** | **Baseline** | **2-28** | **Baseline** | **2-28** | **Baseline** | **2-28** | **Baseline** | **2-28** |
|  | *count* | *count* | *count (%)* | *count (%)* | *count (%)* | *count (%)* | *Mean (SD)* | *Mean (SD)* | *count (%)* | *count (%)* |
| ***Cardiovascular*** |  |  |  |  |  |  |  |  |  |  |
| Venous thromboembolism | 3946 | 17 | 2126 (53.9) | 10 (58.8) | 1820 (46.1) | 7 (41.2) | 67.3 (14.5) | 62.2 (15.8) | 613 (15.5) | <5 |
| Immune thrombocytopenic purpura | 2915 | 15 | 1388 (47.6) | 9 (60.0) | 1527 (52.4) | 6 (40.0) | 63.9 (15.3) | 68.7 (11.9) | 23 (0.8) | 0 (0) |
| Arterial thromboembolism | 9453 | 16 | 4425 (46.8) | 7 (43.8) | 5026 (53.2) | 9 (56.3) | 72.7 (11.9) | 68.3 (12.9) | 1384 (14.6) | <5 |
| Myocardial infarction | 6682 | 13 | 2915 (43.6) | 5 (38.5) | 3766 (56.4) | 8 (61.5) | 72.7 (11.5) | 67.4 (14.2) | 662 (9.9) | <5 |
| Ischaemic stroke | 2975 | 5 | 1605 (53.9) | <5 | 1370 (46.1) | <5 | 74.0 (12.3) | 78.6 (10.8) | 853 (28.7) | <5 |
| Atrial fibrillation | 11575 | 30 | 5682 (49.1) | 15 (50.0) | 5893 (50.9) | 15 (50.0) | 75.1 (11.0) | 69.8 (11.7) | 553 (4.8) | 0 (0) |
| Heart block | 5883 | 11 | 2425 (41.2) | 7 (63.6) | 3458 (58.8) | <5 | 74.2 (12.4) | 63.4 (19.0) | 23 (0.4) | 0 (0) |
| Arrhythmia | 18113 | 62 | 9138 (50.4) | 35 (56.5) | 8975 (49.6) | 27 (43.5) | 71.6 (14.1) | 60.4 (16.9) | 555 (3.1) | 0 (0) |
| ***Neurological*** |  |  |  |  |  |  |  |  |  |  |
| Myasthenia | 766 | 7 | 375 (49.0) | <5 | 391 (51.0) | <5 | 67.6 (16.9) | 55.4 (18.9) | 36 (4.7) | 0 (0) |
| Multiple sclerosis | 3680 | 19 | 2744 (74.6) | 15 (78.9) | 936 (25.4) | <5 | 59.1 (14.4) | 54.4 (9.7) | 205 (5.6) | 0 (0) |
| ***Blood conditions*** |  |  |  |  |  |  |  |  |  |  |
| Aplastic anaemia | 1193 | 9 | 560 (46.9) | <5 | 633 (53.1) | 6 (66.7) | 63.6 (15.6) | 65.4 (21.1) | 28 (2.3) | 1 (11.1) |
| Haemolytic anaemia | 188 | 5 | 100 (53.2) | <5 | 88 (46.8) | <5 | 62.3 (18.9) | 61.4 (15.9) | 9 (4.8) | 0 (0) |
| ***Other conditions*** |  |  |  |  |  |  |  |  |  |  |
| Cholangitis | 954 | 6 | 502 (52.6) | <5 | 452 (47.4) | <5 | 67.4 (15.4) | 54.5 (16.1) | 112 (11.7) | 0 (0) |
| Autoimmune hepatitis | 575 | 5 | 457 (79.5) | <5 | 118 (20.5) | <5 | 60.6 (17.2) | 59.8 (16.2) | 19 (3.3) | 0 (0) |
| Acute renal failure | 15630 | 50 | 7971 (51.0) | 23 (46.0) | 7659 (49.0) | 27 (54.0) | 70.5 (14.4) | 61.4 (16.0) | 356 (2.3) | <5 |
| Acute liver failure | 1425 | 6 | 632 (44.4) | <5 | 793 (55.6) | <5 | 62.5 (13.3) | 47.0 (22.9) | 388 (27.2) | 0 (0) |
| Transplant reject | 936 | 5 | 408 (43.6) | <5 | 528 (56.4) | <5 | 52.3 (16.3) | 49.8 (21.3) | 28 (3.0) | 0 (0) |
| Inflame arthropathy | 5112 | 10 | 3416 (66.8) | 7 (70.0) | 1696 (33.2) | <5 | 69.7 (13.2) | 63.3 (17.2) | 13 (0.3) | 0 (0) |
| Rheumatoid arthritis | 4726 | 23 | 3293 (69.7) | 19 (82.6) | 1433 (30.3) | <5 | 69.0 (13.7) | 57.4 (15.0) | 109 (2.3) | 0 (0) |
| Systemic lupus erythematosus | 771 | 10 | 690 (89.5) | 9 (90.0) | 81 (10.5) | <5 | 55.4 (15.9) | 55.0 (11.8) | 11 (1.4) | 0 (0) |
| Addison's disease | 1132 | 5 | 676 (59.7) | <5 | 456 (40.3) | <5 | 60.9 (16.9) | 66.6 (9.9) | 10 (0.9) | <5 |
| Crohn's disease | 2087 | 9 | 1187 (56.9) | 7 (77.8) | 900 (43.1) | <5 | 49.5 (18.4) | 35.8 (8.0) | 22 (1.1) | 0 (0) |
| Colitis | 2313 | 10 | 1111 (48.0) | 6 (60.0) | 1202 (52.0) | <5 | 55.1 (18.0) | 52.0 (22.0) | 18 (0.8) | 0 (0) |
| Vasculitis | 1678 | 6 | 1086 (64.7) | 5 (83.3) | 1086 (64.7) | 5 (83.3) | 69.9 (15.4) | 66.8 (11.6) | 41 (2.4) | 0 (0) |
| GI bleeding | 5470 | 16 | 2787 (51.0) | 8 (50.0) | 2683 (49.0) | 8 (50.0) | 66.5 (15.7) | 63.5 (16.5) | 366 (6.7) | 0 (0) |

**Supplementary Table 4: Incidence rate ratios (IRR 95% CI) for individual outcomes in pre-defined risk periods immediately before and after treatment, adjusted for calendar time, vaccine status and SARS-CoV-2 positive test from December 11, 2021 until May 24, 2022, SARS-CoV-2 positive test and vaccine status. The total population includes 1,251,852 patients eligible to or treated with nMAB of which 22,815 were treated with Sotrovimab in the study period. Out of 49 outcomes screened for safety signal, only two have showed a significant association with the Sotrovimab treatment (cells with less than 5 events are suppressed)**

|  | **Sotrovimab** | |
| --- | --- | --- |
|  | events | IRR (95% CI) |
| **Rheumatoid arthritis** |  |  |
| Baseline | 4726 | 1.00 |
| -28-1 days | 37 | 1.33 (0.86, 2.05) |
| 0 day | <5 | 0.87 (0.21, 3.63) |
| 1 day | 35 | 19.07 (11.61, 31.32) |
| 2-3 days | 9 | 3.08 (1.44, 6.58) |
| 4-7 days | <5 | 0.63 (0.19, 2.02) |
| 8-28 days | 11 | 0.54 (0.28, 1.04) |
| **Systemic lupus erythematosus** |  |  |
| Baseline | 771 | 1.00 |
| -28-1 days | 6 | 1.39 (0.49, 3.94) |
| 0 day | <5 | n/a |
| 1 day | 5 | 6.06 (1.94, 18.96) |
| 2-3 days | 5 | 5.15 (1.60, 16.60) |
| 4-7 days | <5 | n/a |
| 8-28 days | 5 | 1.50 (0.50, 4.47) |

**Supplementary Table 5: Incidence rate ratios (IRR 95% CI) for individual outcomes in pre-defined risk periods immediately before and after nMAB treatment, adjusted for calendar time, vaccine status and SARS-CoV-2 positive test from December 11, 2021 until May 24, 2022, SARS-CoV-2 positive test and vaccine status. Two models are considered: including only SARS-CoV-2 positive patients; including only SARS-CoV-2 symptomatic patients (cells with less than 5 events are suppressed).**

|  | **Only SARS-CoV-2 positive** | | **Only SARS-CoV-2 symptomatic** | |  | **Only SARS-CoV-2 positive** | | **Only SARS-CoV-2 symptomatic** | |
| --- | --- | --- | --- | --- | --- | --- | --- | --- | --- |
|  | *events* | *IRR (95% CI)* | *events* | *IRR (95% CI)* |  | *events* | *IRR (95% CI)* | *events* | *IRR (95% CI)* |
| **Venous thromboembolism** |  |  |  |  | **Autoimmune hepatitis** |  |  |  |  |
| Baseline | 409 | 1.00 | 160 | 1.00 | Baseline | 94 | 1.00 | 40 | 1.00 |
| -28-1 days | 14 | 1.23 (0.64, 2.40) | <5 | 0.73 (0.19, 2.80) | -28-1 days | 7 | 1.83 (0.63, 5.34) | <5 | 1.45 (0.23, 9.15) |
| 0 day | <5 | n/a | <5 | n/a | 0 day | <5 | 8.98 (1.68, 47.89) | <5 | n/a |
| 1 day | <5 | 3.43 (1.04, 11.33) | <5 | n/a | 1 day | 5 | 44.41 (13.58, 145.28) | <5 | 108.09 (15.18, 769.83) |
| 2-28 days | 17 | 0.76 (0.42, 1.41) | 6 | 0.58 (0.20, 1.67) | 2-28 days | 5 | 1.95 (0.60, 6.30) | <5 | 2.81 (0.44, 17.94) |
| **Immune thrombocytopenic purpura** |  |  |  |  | **Acute renal failure** |  |  |  |  |
| Baseline | 258 | 1.00 | 103 | 1.00 | Baseline | 1369 | 1.00 | 462 | 1.00 |
| -28-1 days | 18 | 1.12 (0.60, 2.12) | <5 | 0.87 (0.17, 4.61) | -28-1 days | 39 | 0.93 (0.63, 1.36) | 8 | 0.55 (0.25, 1.22) |
| 0 day | <5 | n/a | <5 | n/a | 0 day | <5 | 0.68 (0.17, 2.78) | <5 | n/a |
| 1 day | <5 | n/a | <5 | n/a | 1 day | 9 | 3.17 (1.59, 6.30) | <5 | 1.96 (0.47, 8.22) |
| 2-28 days | 14 | 0.79 (0.40, 1.54) | 9 | 1.38 (0.51, 3.77) | 2-28 days | 47 | 0.74 (0.52, 1.06) | 29 | 1.19 (0.71, 1.99) |
| **Arterial thromboembolism** |  |  |  |  | **Acute liver failure** |  |  |  |  |
| Baseline | 760 | 1.00 | 236 | 1.00 | Baseline | 76 | 1.00 | 31 | 1.00 |
| -28-1 days | 19 | 1.17 (0.67, 2.04) | 7 | 2.04 (0.76, 5.47) | -28-1 days | <5 | 1.42 (0.31, 6.44) | <5 | 2.23 (0.27, 18.46) |
| 0 day | <5 | n/a | <5 | n/a | 0 day | <5 | n/a | <5 | n/a |
| 1 day | 9 | 10.75 (5.18, 22.32) | <5 | n/a | 1 day | <5 | n/a | <5 | n/a |
| 2-28 days | 15 | 0.81 (0.44, 1.48) | 7 | 1.34 (0.51, 3.51) | 2-28 days | 6 | 2.04 (0.57, 7.36) | <5 | 1.78 (0.22, 14.18) |
| **Myocardial infarction** |  |  |  |  | **Transplant reject** |  |  |  |  |
| Baseline | 546 | 1.00 | 169 | 1.00 | Baseline | 107 | 1.00 | 45 | 1.00 |
| -28-1 days | 14 | 0.98 (0.51, 1.85) | 5 | 1.25 (0.41, 3.86) | -28-1 days | <5 | 1.02 (0.29, 3.55) | <5 | n/a |
| 0 day | <5 | n/a | <5 | n/a | 0 day | <5 | n/a | <5 | n/a |
| 1 day | 6 | 8.64 (3.56, 20.96) | <5 | n/a | 1 day | <5 | 5.95 (1.63, 21.72) | <5 | n/a |
| 2-28 days | 13 | 0.89 (0.46, 1.72) | 6 | 1.09 (0.38, 3.11) | 2-28 days | 5 | 0.40 (0.14, 1.18) | <5 | n/a |
| **Ischaemic stroke** |  |  |  |  | **Inflame arthropathy** |  |  |  |  |
| Baseline | 225 | 1.00 | 67 | 1.00 | Baseline | 524 | 1.00 | 185 | 1.00 |
| -28-1 days | <5 | 1.33 (0.39, 4.51) | <5 | 2.26 (0.34, 15.19) | -28-1 days | 16 | 1.13 (0.62, 2.05) | <5 | 0.59 (0.16, 2.14) |
| 0 day | <5 | n/a | <5 | n/a | 0 day | <5 | n/a | <5 | n/a |
| 1 day | <5 | n/a | <5 | n/a | 1 day | 11 | 15.35 (7.75, 30.40) | <5 | 11.79 (3.82, 36.42) |
| 2-28 days | <5 | 0.71 (0.21, 2.37) | <5 | 1.46 (0.29, 7.36) | 2-28 days | 9 | 0.54 (0.26, 1.12) | 6 | 0.73 (0.28, 1.92) |
| **Atrial fibrillation** |  |  |  |  | **Rheumatoid arthritis** |  |  |  |  |
| Baseline | 1020 | 1.00 | 340 | 1.00 | Baseline | 541 | 1.00 | 214 | 1.00 |
| -28-1 days | 19 | 0.58 (0.35, 0.97) | <5 | 0.23 (0.07, 0.77) | -28-1 days | 34 | 1.29 (0.82, 2.03) | 7 | 0.59 (0.24, 1.45) |
| 0 day | <5 | n/a | <5 | n/a | 0 day | <5 | n/a | <5 | n/a |
| 1 day | 7 | 4.46 (2.04, 9.78) | <5 | n/a | 1 day | 34 | 25.70 (16.33, 40.47) | 14 | 17.42 (8.65, 35.09) |
| 2-28 days | 30 | 0.84 (0.54, 1.31) | 11 | 0.64 (0.31, 1.32) | 2-28 days | 21 | 0.68 (0.41, 1.15) | 9 | 0.45 (0.20, 0.99) |
| **Heart block** |  |  |  |  | **Systemic lupus erythematosus** |  |  |  |  |
| Baseline | 567 | 1.00 | 185 | 1.00 | Baseline | 122 | 1.00 | 54 | 1.00 |
| -28-1 days | 16 | 0.83 (0.46, 1.52) | 5 | 1.17 (0.38, 3.54) | -28-1 days | 5 | 1.37 (0.44, 4.31) | <5 | n/a |
| 0 day | <5 | n/a | <5 | n/a | 0 day | <5 | n/a | <5 | n/a |
| 1 day | <5 | 4.94 (1.74, 14.00) | <5 | 8.39 (1.80, 39.17) | 1 day | 5 | 14.17 (4.82, 41.70) | <5 | 13.79 (2.44, 77.87) |
| 2-28 days | 11 | 0.56 (0.28, 1.12) | 5 | 0.87 (0.29, 2.59) | 2-28 days | 10 | 1.35 (0.56, 3.24) | 5 | 1.55 (0.41, 5.84) |
| **Arrhythmia** |  |  |  |  | **Addison's disease** |  |  |  |  |
| Baseline | 1789 | 1.00 | 599 | 1.00 | Baseline |  |  | 53 | 1.00 |
| -28-1 days | 44 | 0.79 (0.56, 1.12) | 15 | 1.11 (0.60, 2.06) | -28-1 days |  |  | <5 | n/a |
| 0 day | <5 | n/a | <5 | n/a | 0 day |  |  | <5 | n/a |
| 1 day | 12 | 4.17 (2.30, 7.57) | <5 | 2.00 (0.48, 8.34) | 1 day |  |  | <5 | n/a |
| 2-28 days | 61 | 0.91 (0.67, 1.25) | 21 | 0.90 (0.53, 1.55) | 2-28 days |  |  | <5 | n/a |
| **Myasthenia** |  |  |  |  | **Crohn's disease** |  |  |  |  |
| Baseline | 92 | 1.00 | 29 | 1.00 | Baseline | 449 | 1.00 | 202 | 1.00 |
| -28-1 days | <5 | 1.05 (0.24, 4.53) | <5 | n/a | -28-1 days | 10 | 0.84 (0.41, 1.73) | <5 | 0.36 (0.08, 1.61) |
| 0 day | <5 | n/a | <5 | n/a | 0 day | <5 | n/a | <5 | n/a |
| 1 day | <5 | 26.05 (6.49, 104.59) | <5 | 38.85 (4.80, 314.44) | 1 day | 14 | 27.35 (14.32, 52.25) | <5 | 12.19 (3.46, 42.95) |
| 2-28 days | 7 | 2.39 (0.70, 8.16) | <5 | 1.21 (0.17, 8.72) | 2-28 days | 8 | 0.64 (0.29, 1.41) | <5 | 0.63 (0.21, 1.93) |
| **Multiple sclerosis** |  |  |  |  | **Colitis** |  |  |  |  |
| Baseline | 516 | 1.00 | 187 | 1.00 | Baseline | 395 | 1.00 | 156 | 1.00 |
| -28-1 days | 21 | 1.41 (0.80, 2.48) | 11 | 2.56 (1.06, 6.15) | -28-1 days | 8 | 0.93 (0.41, 2.13) | <5 | 0.94 (0.19, 4.57) |
| 0 day | 5 | 3.33 (1.25, 8.83) | <5 | 6.24 (1.36, 28.54) | 0 day | <5 | n/a | <5 | n/a |
| 1 day | 36 | 40.25 (24.73, 65.52) | 12 | 36.07 (15.79, 82.38) | 1 day | <5 | 10.23 (3.50, 29.93) | <5 | n/a |
| 2-28 days | 19 | 0.97 (0.55, 1.73) | 9 | 1.11 (0.45, 2.70) | 2-28 days | 10 | 1.10 (0.51, 2.34) | 7 | 1.70 (0.62, 4.70) |
| **Aplastic anaemia** |  |  |  |  | **Vasculitis** |  |  |  |  |
| Baseline | 111 | 1.00 | 45 | 1.00 | Baseline | 179 | 1.00 | 68 | 1.00 |
| -28-1 days | 11 | 2.42 (0.96, 6.10) | <5 | 0.78 (0.15, 3.90) | -28-1 days | 7 | 0.99 (0.38, 2.55) | <5 | n/a |
| 0 day | <5 | n/a | <5 | n/a | 0 day | <5 | n/a | <5 | n/a |
| 1 day | <5 | 6.66 (1.42, 31.20) | <5 | n/a | 1 day | 11 | 35.13 (15.21, 81.11) | <5 | 60.57 (13.87, 264.55) |
| 2-28 days | 8 | 1.18 (0.45, 3.12) | <5 | 0.71 (0.14, 3.52) | 2-28 days | 5 | 0.61 (0.22, 1.74) | <5 | 1.82 (0.45, 7.37) |
| **Haemolytic anaemia** |  |  |  |  | **GI bleeding** |  |  |  |  |
| Baseline | 27 | 1.00 | 11 | 1.00 | Baseline | 495 | 1.00 | 161 | 1.00 |
| -28-1 days | <5 | n/a | <5 | n/a | -28-1 days | 15 | 0.93 (0.50, 1.73) | <5 | 0.85 (0.26, 2.77) |
| 0 day | <5 | n/a | <5 | n/a | 0 day | <5 | n/a | <5 | n/a |
| 1 day | <5 | n/a | <5 | n/a | 1 day | <5 | 4.64 (1.42, 15.23) | <5 | 9.62 (2.07, 44.69) |
| 2-28 days | 5 | 0.93 (0.18, 4.82) | <5 | n/a | 2-28 days | 16 | 1.00 (0.55, 1.83) | 5 | 0.92 (0.31, 2.72) |
| **Cholangitis** |  |  |  |  |  |  |  |  |  |
| Baseline | 108 | 1.00 | 33 | 1.00 |  |  |  |  |  |
| -28-1 days | <5 | 0.70 (0.21, 2.36) | <5 | n/a |  |  |  |  |  |
| 0 day | <5 | n/a | <5 | n/a |  |  |  |  |  |
| 1 day | <5 | n/a | <5 | n/a |  |  |  |  |  |
| 2-28 days | 6 | 1.39 (0.46, 4.17) | <5 | 1.59 (0.23, 11.08) |  |  |  |  |  |

**Supplementary Figure 1: Data flow diagram**

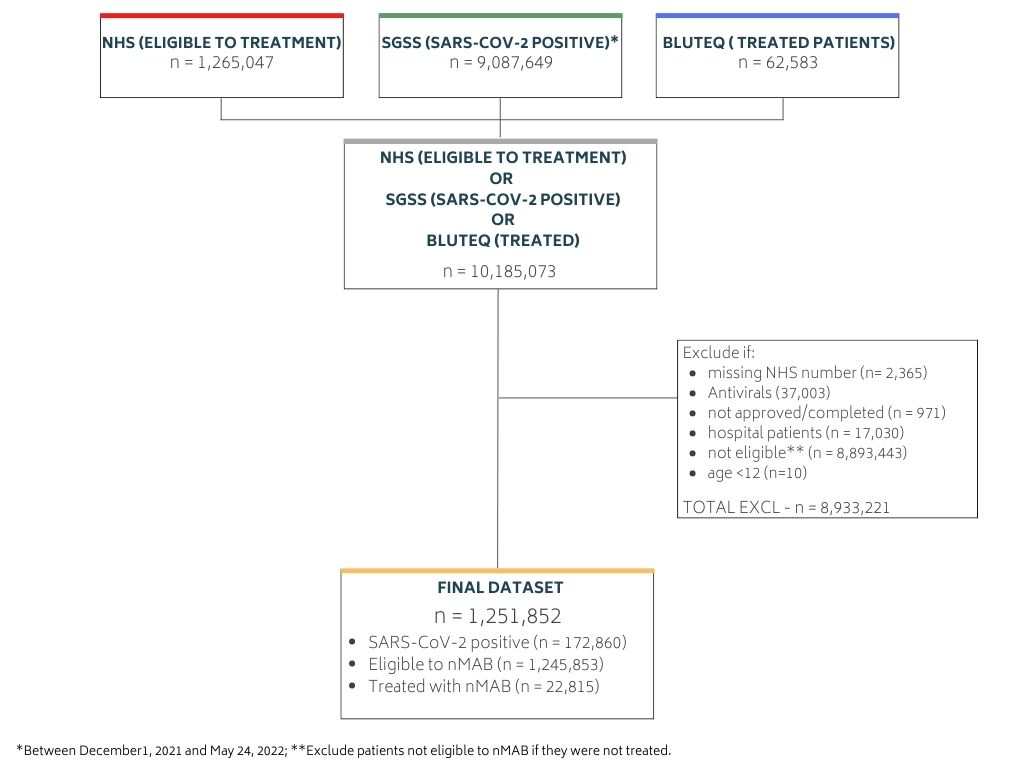
